# Co-creating a *Choosing Wisely* Leaflet Supporting the Reduction of Imaging Usage in Low Back Pain Management - A Multi-Method Study

**DOI:** 10.1101/2024.07.02.24309693

**Authors:** Chris Djurtoft, Edel O’Hagan, Monika D. Laursen, Lars Lejbølle, Mia Bisgaard, Simon K. Johansen, Kristian D. Lyng, Morten Hoegh, Negar Pourbordbari, Malene K. Bruun, Bettina Eiger, Jesper B. Larsen, Michael S. Rathleff

## Abstract

**Introduction:** Low back pain is a global health challenge with negative consequences for both individuals and healthcare systems. The *Choosing Wisely* initiative aims to improve communication between patients and clinicians to promote informed healthcare decisions while avoiding unnecessary tests and treatments. The objective of this project is to co-create an information resource in the form of a leaflet, to be distributed in clinical settings, websites or social media targeting people with low back pain.

**Methods:** This multi-method study was conducted in four stages: literature search, input from practice consultants, program theory development, and think-aloud interviews with people experiencing low back pain. Each stage was followed by a consensus meeting in which the steering group refined the leaflet based on the emerging knowledge.

**Results:** The literature search highlighted patients’ need for understandable information about diagnosis, treatment options, and self-management strategies. Practice consultants emphasized concise, relatable content. The program theory identified potential mechanisms and design targets for content creation, development, and implementation of the leaflet, such as addressing patient concerns, reducing diagnostic uncertainty, offering insights into management options, and validation. Think-aloud interviews with 18 people living with low back pain informed the iteration of the leaflet, enhancing language clarification and content comprehension.

**Conclusion:** We co-created a new *Choosing Wisely* leaflet, created with end-users in mind, specifically focused on reducing unnecessary imaging for low back pain. Valuable feedback from end-users prompted significant revisions, underscoring the importance of involving multiple end-user viewpoints into the creation process.

## Introduction

Low back pain is a significant public health concern, affecting a large portion of the population and causing a considerable impact on individual quality of life, work abilities and participation in activities [18,26,27,35,43]. Although spending on health services has increased, people seeking care for their low back pain still receive services known to provide low-value or potentially cause harm [4]. These low-value care options include unnecessary diagnostic imaging,[28] opioids,[1,48] antidepressants,[17] muscle relaxants,[6] and spinal fusion surgery [24]. This, despite international clinical guidelines finding insufficient evidence for these management options [30,42,62].

An evaluation of routine data collected from patients that attended primary care, showed that fewer than 5% of the magnetic resonance imaging (MRI) requested were clearly indicated [53]. This finding coupled with the growing concern of unnecessary imaging for low back pain underscores the need for strategies to improve the accuracy of imaging decisions [30,31]. The mechanisms involved are complex and may illustrate the multifaceted impact of diagnostic uncertainty, fear of making mistakes (such as defensive medicine), and patient pressure on clinicians’ decision-making processes during consultations [2,10,30,56]. Clinicians’ ability to handle diagnostic uncertainty and defensive medicine holds significance across various domains; this affects decision-making and may result in an increased number of referrals to specialized sectors and medical tests [3,8–10,55].

Patient informational resources have demonstrated efficacy in facilitating informed decision-making and improving health outcomes compared to usual care [19,59]. These materials can significantly improve health knowledge, accurate risk perceptions, and patients’ active participation in decision-making [19,59]. Certain types of information prove more influential in shaping health behaviors in people living with low back pain [19,49]. Targeted information focusing on the cause and severity of low back pain has the potential to diminish obstacles to physical activity and improve their activity levels [19,49]. However, Insufficient reporting on material development limits the understanding of the effectiveness and key mechanism of the current informational resources [19,59].

*Choosing Wisely* is a healthcare campaign aimed at encouraging healthcare professionals and patients to make informed and evidence-based decisions about medical tests and treatments [23,40]. It promotes the idea of using healthcare resources wisely by avoiding unnecessary or low-value care and facilitate evidence based-decision making [40]. Previous *Choosing Wisely* campaigns have shown mixed effectiveness, in terms of clinical decision support and imaging usage [7]; most interventions focus solely on clinicians, despite that imaging requests frequently originate from patients [23,41,55]. This emphasizes the importance of targeting both patients and clinicians in co-creation processes to ensure that the materials meet their needs and address key outcomes, such as information relevance, clarity, and patient satisfaction [45,57,58].

The objective of this study is to co-create an information resource in the form of a leaflet that can be distributed in clinical settings, websites, and social media, targeting individuals experiencing low back pain. The purpose of the leaflet is to serve as a conversation tool between patients, relatives, and clinicians during consultations, aiming to reduce the use of imaging diagnostics (MRI or X-ray), when there is no clinical suspicion of serious illness.

## Methods

This project was led by a steering group (CD, LL, MDL, MB, MSR) with input from a project group (EO, KDL, MH, JBL, MKB, BE, NP). Additionally, the steering group received ongoing input from practice consultants (licensed healthcare professionals currently working with people experiencing low back pain in clinical practice) in the North Denmark Region throughout the process. This multiple-method study employed an iterative process conducted in four stages (Figure 1):

1. Systematic literature search aimed to answer; *“which information needs to be included in a leaflet for people experiencing low back pain?”,*
2. Input from practice consultants to explore: *“the needs of clinical practice, what is considered important in a leaflet, and what information should be covered?”*,
3. Development of a program theory to support the *Choosing Wisely* leaflet, and co-creation of the information resource (*Choosing Wisely* leaflet),
4. Think-aloud interviews with adults experiencing low back pain that aimed to understand end-users’ perspectives regarding the leaflet.

**Figure 1.**
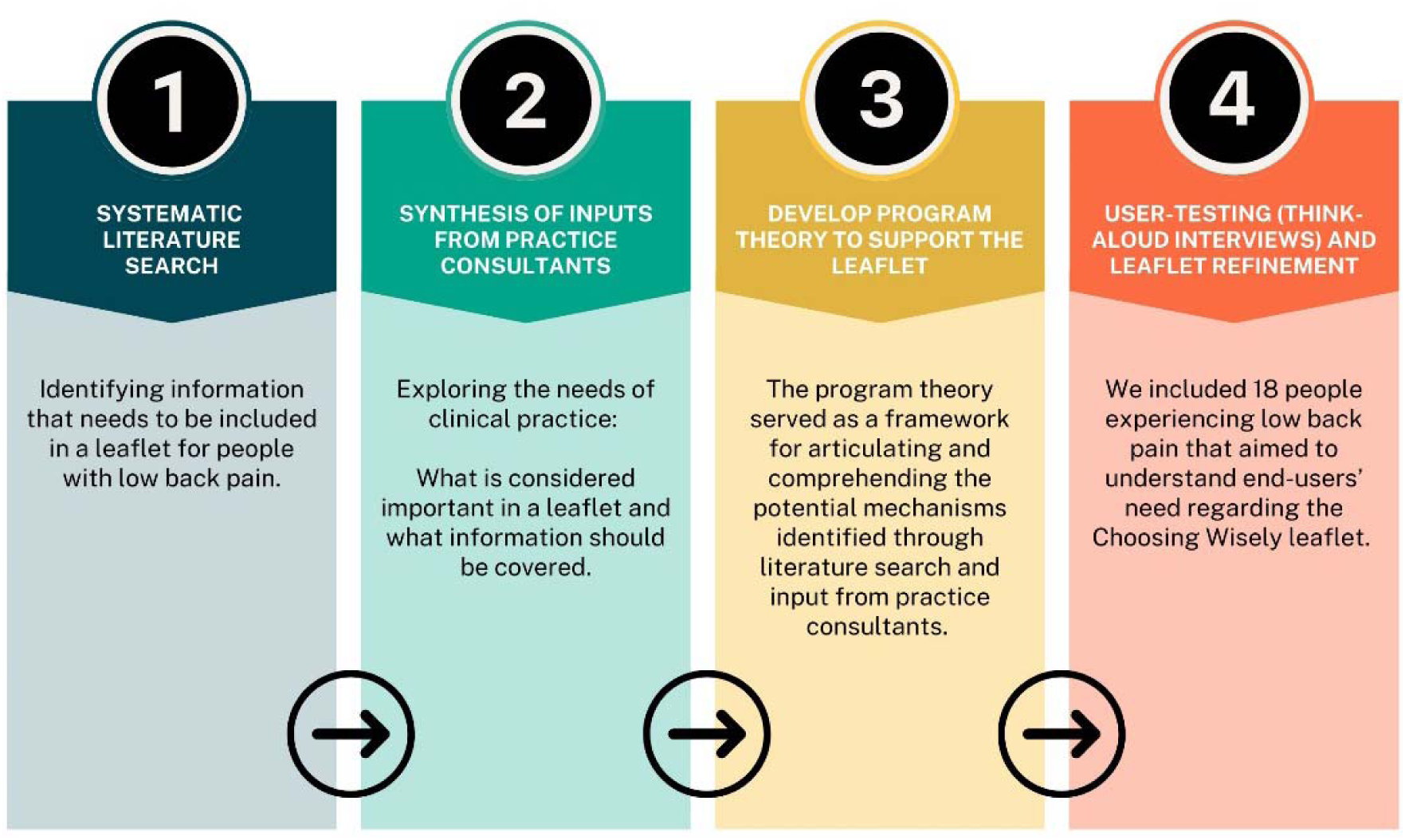
The iterative process conducted in four stages. This multiple-method study used an iterative process conducted in four stages, followed by consensus meetings within the steering group with representation of members from the project group.

Each of the four stages was followed by consensus meetings in the steering group with representation of members from the project group, which through an iterative process developed and designed the leaflet based on the generated knowledge. North Denmark Region Committee on Health Research Ethics determined that this study was exempt from ethical approval because of its non-interventional design. Signed informed consent was obtained from all participants during the think-aloud interviews. All data were stored on a local secure fileshare hosted at Aalborg University and the North Denmark Region.

### Stage 1 - Synthesis of inputs from the literature search

#### Search strategy and study eligibility

In May 2023, three researchers (CD, BE, and MSR) independently identified studies (qualitative and quantitative) published between 2000 and May 2023. Eligible studies were those focusing on information needed by adults to understand low back pain, defined according to the chronic primary musculoskeletal pain criteria outlined by Nicholas et al 2019 [47]. We searched MEDLINE (via PubMed). The search strategy was initially designed by CD in collaboration with MSR, and revisions and edits to the content were provided before being validated among all members in the project group. Search terms related to education and information needs were combined with low back pain in the search strategy (appendix 1).

#### Data analysis

Relevant references were exported to Microsoft Excel 365 via PubMed, before independently selected (title/abstract) by CD, BE, and MSR. We used information from all included articles, such as qualitative statements and quantitative findings, to form the basis for the content and structure of the programme theory and *Choosing Wisely* leaflet. We summarized the main overall findings in a brief narrative summary that was discussed and agreed upon at a consensus meeting within the project group before developing the programme theory (appendix 2).

### Stage 2 – Synthesis of inputs from practice consultants

The aim of this stage was to explore the needs and preferences from clinical practice. We included (n=5) practice consultants (a general practitioner, a physiotherapist, a chiropractor, a rheumatologist, and a neurosurgeon) that provided input to the development of the *Choosing Wisely* leaflet based on their clinical expertise. These consultants were recruited from general practitioner clinics and medical networks in the North Denmark Region to ensure diverse representation. Their involvement was facilitated through written email communication including questions such as “How should a leaflet look to assist your clinical practice?”. Additionally, some shared references to illustrate their point [15].

#### Data analysis

The practice consultants’ responses were extracted into Microsoft Word 365. Each response was read by JBL and MKB, and findings were synthesized into a narrative summary. This summary was then discussed and agreed upon during a consensus meeting with all involved parties (appendix 3). These detailed perspectives were used to inform the refinement and improvement of the *Choosing Wisely* leaflet’s content and usability.

### Stage 3 - Development of a program theory to support the *Choosing Wisely* leaflet

We used the findings from the literature search and synthesis of inputs from practice consultants to construct a program theory. The program theory served as a framework for articulating and comprehending the underlying mechanisms identified through literature search and input from practice consultants and was structured in six subheadings as described by Kazi et al [36]: Target group (context); Activities; Effective mechanisms (mechanisms); Short-term outcome; Medium-term outcome; Long-term outcomes (appendix 4). MSR and SKJ synthesized the findings into a structured programme theory before presenting it to the steering and project group. The theory was subsequently iteratively agreed upon at a consensus meeting. This process ensured that the program theory accurately reflected the synthesized information and received input and endorsement from relevant stakeholders before progressing further.

We created the initial draft of the *Choosing Wisely* leaflet using the identified potential pathways and mechanisms expected to drive the leaflet’s intended outcomes. Additionally, we extracted information from existing leaflets found through searches on PubMed, Google, and Google Scholar [23,31]. The content was designed using a second person perspective, employing simple language and an informal tone, such as “it is rarely necessary for you to undergo scanning…”, to boost relatability. Any disparities were addressed through discussions and reached a consensus among the members in the steering and project group prior to initiating stage 4.

### Stage 4 - Think-aloud interviews (User testing) and refinement of the *Choosing Wisely* leaflet

We employed individual qualitative think-aloud interviews, to actively engage end-users in the development process through user testing [5,29]. The think-aloud method offers a simulation-based approach to understanding end-users’ perceptions, interactions, reasoning processes, and latent challenges associated with using the *Choosing Wisely* leaflet [5,29]. This qualitative design approach was divided into four separate sessions to ensure iterative refinement of the end-users’ perspectives, aligning it with the needs and preferences of people experiencing low back pain. It involved iterative refinement of the design based on the acquired knowledge [5,29]. All demographic data was collected via Research Electronic Data Capture (REDCap^TM^) [25].

#### End-user involvement: People experiencing low back pain

Participants experiencing low back pain were recruited by convenience sampling using the practice consultant’ clinics in the North Denmark Region, and social media posts via Facebook. We included adults experiencing low back pain, ≥18 years old, and able to speak and read Danish. We decided to divide the interviews into separate sessions with at least five adults in each session to ensure a variety of perspectives [5,29]. We included six participants for each of the four sessions to achieve sufficient information power in the iterations [16,44]. The interview guide was first pilot tested internally within the author group, and subsequently with two individuals experiencing low back pain prior to the first interview (appendix 5) [33]. Prior to the interview participants with low back pain completed a demographic questionnaire, including items on gender, age, current low back pain (yes/no), physical activity, pain medication, low back pain duration, and pain in other body locations (appendix 6). All interviews took place online via Microsoft Teams and were video recorded. Participants were instructed to describe their thoughts as they occurred, saying what came to mind as they went through the *Choosing Wisely* leaflet [5]. MDL reminded the participants to keep talking if they fell silent, by using prompts such as *“What are you thinking of right now“*, or *“You are not saying anything right now?“*. The aim of the think-aloud interviews was to pinpoint any disparities between the participants’ interpretation of the text and its intended meaning. Additionally, MDL gathered field notes during the interviews that were later refined by reviewing the video recordings to ensure that emerging, relevant insights were identified post-hoc and addressed in the following interviews [5,29].

#### Data analysis – Iterative revisions

All changes to the *Choosing Wisely* leaflet was based on re-watching the video, notes, and transcriptions. All online interviews were transcribed non-verbatim to retain their meaning, as our goal was to focus on identifying comprehension issues as well as additional suggestions, and we did not plan to undertake an extensive thematic analysis [22,37]. After the first think-aloud session, revisions were collectively decided upon during a steering group meeting, informed by interview data, prior to moving on to the next think-aloud session. This iterative process continued until suggestions for revisions were minimized, as recommended [5,29]. The practice consultants had the opportunity to provide feedback before the first session and after the last session. The insights gained from the previous steps informed the design choices made during the modulation of the leaflet in each round. This ensured that the final leaflet was both user-friendly and aligned with the needs identified through program theory, user feedback, and expert input.

## Results

### Summary of the systematic search

The literature search emphasized that individuals with low back pain need clear and consistent information about the diagnosis, treatment options, prognosis, and self-management strategies, delivered in calming and reassuring terms (appendix 2).

### Synthesis of the practice consultants

The practice consultants emphasized a focus on providing understandable information about low back pain, self-management strategies, and treatment options to reduce the proportion of MRI scans. It was recommended that the leaflet targeted people who were at risk of poor prognosis, such as having negative thoughts about low back pain and its management. Furthermore, it was highlighted that the leaflet should be designed to be distributed by healthcare professionals with additional explanations, made available online, and possibly supplemented with QR codes linking to other materials, such as informational videos. Lastly, it was emphasized that the leaflet was kept concise, with a maximum of two pages, to ensure easy comprehension of key points and to fit within the consultation.

### Program theory and development of the Choosing Wisely leaflet

The program theory identified potential mechanisms guiding the structure, development, and implementation of the leaflet. This involved addressing patient concerns, reducing diagnostic uncertainty, providing decision support, offering insights into management options, and overcoming barriers such as patient insistence on imaging diagnostics. Short-term outcomes included reduced diagnostic uncertainty, patient reassurance, reduction of anxiety, and validating the experience, while medium-term outcomes focused on reducing both diagnostic uncertainty and physician’s fear of making mistakes, often leading to the practice of defensive medicine. Long-term outcomes encompassed decreased unnecessary imaging diagnostics, cost reduction, increased patient satisfaction, and clinicians improved understanding of integrating informational materials into patient care.

### Stage 4 – End-user testing: Think-aloud interviews

We included 18 people experiencing low back pain (six in each session). The median age of participants was 37, ranging from 20 to 80 years, and 83% were female (Table 1). The duration of the interviews ranged from 25 to 39 minutes, with a mean duration of 32 minutes.

**Table 1:**
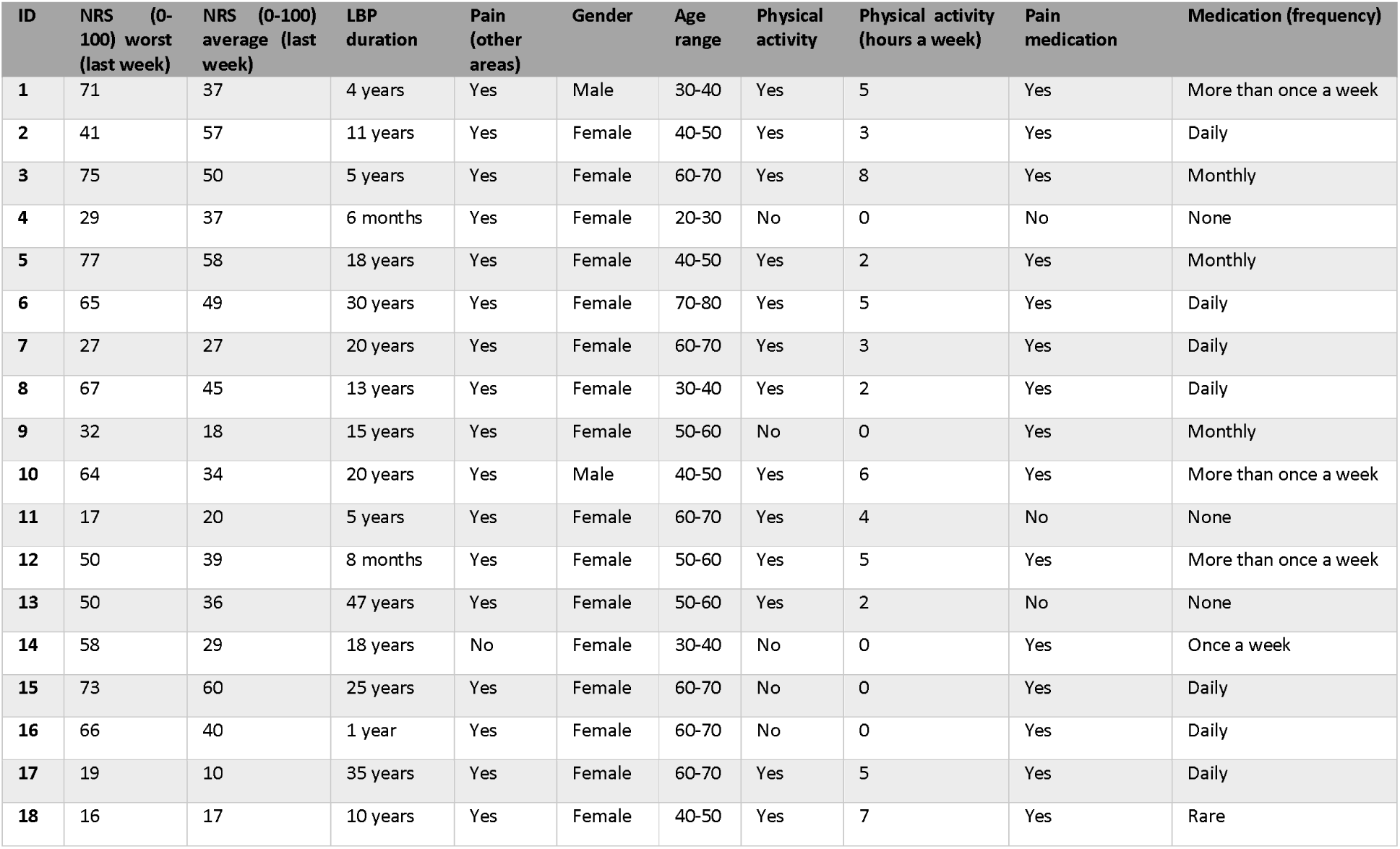
Demographic characteristics. NRS (0-100) worst (last week): Numeric Rating Scale score representing the worst pain experienced by participants in the past week (0 = no pain, 100 = worst imaginable pain). NRS (0-100) average (last week): Numeric Rating Scale score representing the average pain experienced by participants in the past week (0 = no pain, 100 = worst imaginable pain). Pain (other areas): presence of pain in areas other than the lower back. Abbreviation: NRS = Numeric Rating Scale. LBP = Low back pain. See appendix 6 for full Research Electronic Data Capture (REDCap) questionnaire.

Most of the changes to the leaflet were made following the first two think-aloud sessions. The primary issues were the use of improper language, such as medical terminology, the need for clarification of content, and unclear phrasing. This improved the layout and usability with simplified language, visual illustrations, and a focus on practical solutions adjusted to manage low back pain. In the following sessions, although the reasons for changes remained similar, comprehension issues decreased, while participants noted an enhanced reading flow. The participants also appreciated the relatable content, such as validation of their experiences, concerns, and pain-related worries, which reflected their own lived experiences:

> *“I think it is really nice, this here. I must say […] because I think they’re trying to calm down your alarm bells here and guide you a bit, which I think is really nice. It’s also something that [my physiotherapist] has worked a lot on with me, ‘calm down, we’ll get it under control.’ So it could well be that we made this one here”.*

Participants mentioned that it was important that the explanations provided reassurance and was easy to read:

> *”It’s extremely easy to read, very accessible […] I would almost say that if I read it the first time I had back pain, it would be somewhat reassuring, like ‘okay, this isn’t something dangerous, many people have this.’ That’s the immediate impression I get from it”.*

This led us to simplify the language and add more explanatory illustrations to make the content more accessible. It was highlighted that frustration often accompanies dealing with challenging situations, particularly when it involves uncertainty of causes and not knowing the trajectory of low back pain:

> *”The easiest thing you can do is to start feeling frustrated when you’re dealing with something like that [low back pain]. Partly to get it described and so on […] For example, if it was hanging at the doctor’s office when I got there, I would grab it right away”.*

The participants emphasized the importance of information being concise, easy to read, and free of technical/medical terms, so that it was accessible and understandable without missing key points. Too much text or complicated words could discourage the reader from engaging with the content:

> *”It is not crammed with too much stuff where you think, ‘Oh, I need to read that again because I didn’t understand it at all.’ There aren’t a lot of technical terms or anything like that, so I think it is extremely easy to read, it is very accessible. And it’s not too long either, so you don’t only get halfway through and then can’t read the rest.”*

#### Refinement and co-creation of the Choosing Wisely leaflet

The *Choosing Wisely* leaflet was developed over the course of six status meetings in the steering group and subsequently approved among all members in the project group (Figure 2). In total, we made 95 changes as a result of input from think-aloud interviews from the first version of the leaflet to the last version.

**Figure 2.**
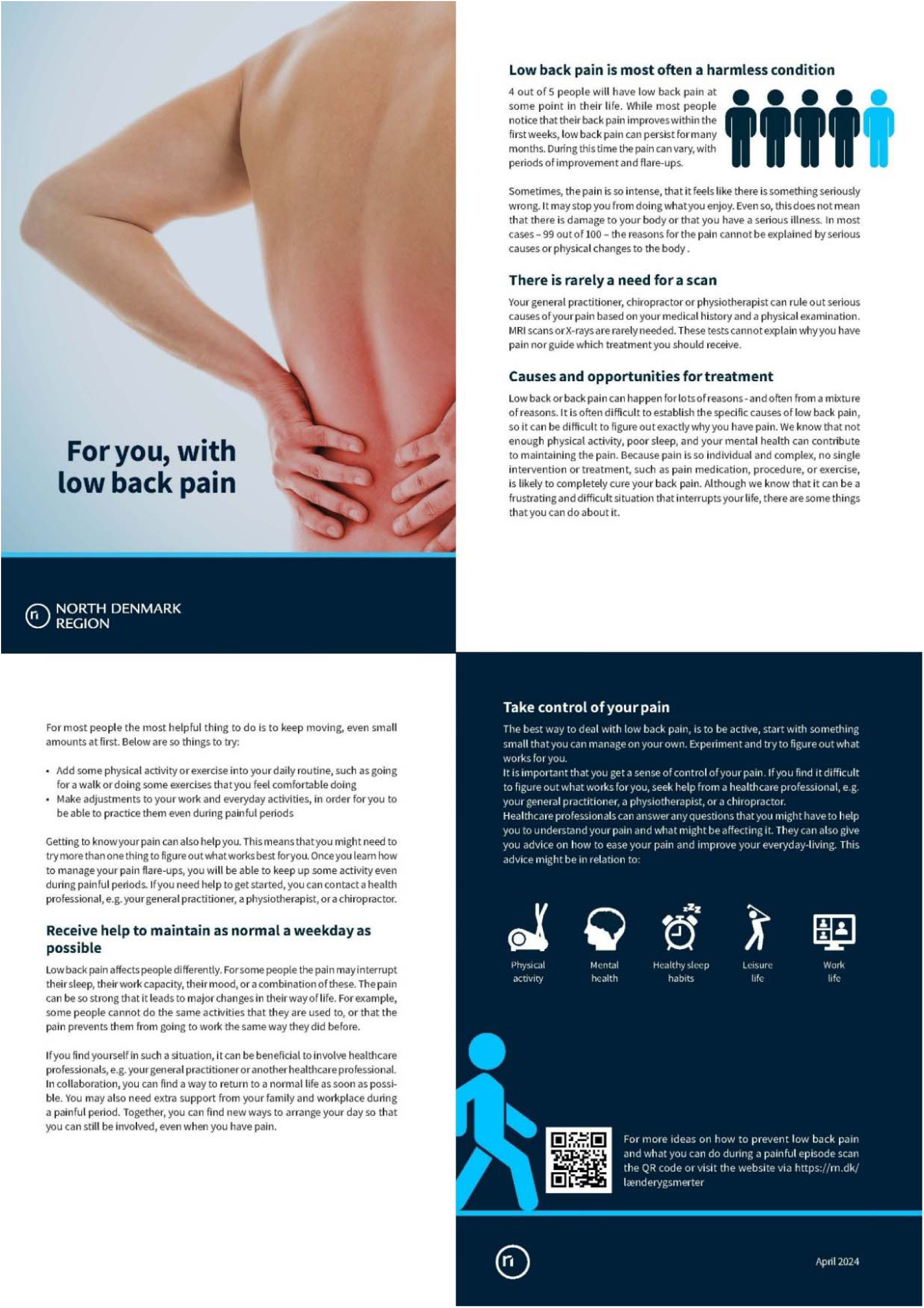
The leaflet is on an A4 page that is folded in the middle. The English version of the *Choosing Wisely* leaflet have not been think aloud tested. Only the Danish version have been revised based on end-user perspectives (Think-aloud interviews). Therefore, the English version of the *Choosing Wisely* leaflet is our translation exclusively.

The leaflet consists of four sections allocated on an A4 page that is folded in the middle. It includes figures and various colours to illustrate the content. Additionally, the leaflet contains a QR code that links to a webpage with online/digital versions of the leaflet. This webpage includes detailed information and needs regarding low back pain that were identified during the think aloud interviews, supplementing the content of the *Choosing Wisely* leaflet.

## Discussion

We co-created a new *Choosing Wisely* leaflet aimed at reducing the use of imaging diagnostics in people with low back pain. Our multi-method study used a program theory to shape the content and design, and we included end-users to enhance co-creation. The initial version reflected what clinicians and researchers deemed necessary according to the program theory. However, through iterative user-testing with individuals who have lived experience of low back pain, the leaflet underwent significant changes in needs, appearance, and phrasing. This highlighted the importance of balancing theoretical theories with end-user involvement to create user-friendly health communication materials [13,20,29].

### Explanations of findings

#### Involving end users to co-create the Choosing Wisely leaflet

End-user engagement throughout the process facilitated the identification and mitigation of prevalent misconceptions regarding low back pain. Recent findings have emphasized the importance individuals place on understanding the causes of pain, as it may influence perceived management strategies, and needs for additional examinations [8,11,38,61]. Sharma et al. found that both patients and clinicians consider imaging useful for identifying the source of non-specific low back pain; patients viewing pathological findings as validation of their pain’s legitimacy [55]. Patients often seek imaging for low back pain due to the belief that it will diagnose their condition; however, they may not fully grasp the risks and harms associated with unnecessary imaging, such as increased uncertainty and invalidation of their pain [3,8,52,55]. It appears that people with low back pain resort to social media for validation [50], suggesting that clinician advice or explanations might not be resonating as intended [54]. Distilling these types of messages into scalable resources that can be widely distributed seems necessary to counteract the prevalent unhelpful messages about low back pain and reduce diagnostic uncertainty [30,31,55]. We implemented 95 modifications during the user-test which emphasizes the challenge and importance of developing structured health information aligned with the perceived needs and comprehension of the target audience.

#### Defensive medicine

Defensive medicine is widespread and needs system wide changes. The drivers of defensive medicine are complex and include both assurance and avoidance behaviors; these practices have significant implications for healthcare systems, such as increased costs and potential harms through overdiagnosis and overtreatment [32,51,52]. This includes unnecessary tests or procedures to reassure patients or protect against the risk of legal action [32,51,52]. Clinicians may feel pressured by patients or their relatives to order imaging, particularly when they lack confidence or experience, or when they wish to meet the expectations [3,52]. However, clinical factors such as patient-clinician collaboration may contribute to reduced overuse of imaging [3,52]. This combination of patient expectations and systemic or organizational pressures can lead to unnecessary imaging that may not improve patient outcomes [12,21,30,52]. The informational *Choosing Wisely* leaflet has the potential to align patient expectations and clinician beliefs regarding appropriate low back pain care, potentially mitigating defensive medicine practices [30,39]. Future studies should investigate the leaflet’s effectiveness on imaging utilization rates.Implementing evidence-based strategies

Healthcare systems need to adopt strategies that enhance implementation of evidence-based practices and guidelines to reduce treatment overuse [3,21,52]. However, implementing guidelines in complex healthcare settings is challenging because of its multiple implications, such as individual patient needs, administrative processes, and the different clinical environments [46]. Effective implementation, particularly clinician-focused and multicomponent strategies, is crucial for integrating *Choosing Wisely* guidelines into clinical settings [7]. Despite the benefits of informational resources, usage in clinical practice is low due to barriers, such as lack of funding, implementation mechanisms (i.e., context-specific needs), and support from healthcare professionals/settings [19,59]. Key facilitators include online/digital delivery, co-creation with patients, cross-sectorial collaborations, clinician training and awareness, and leadership support [19,59,60]. Therefore, we gathered a cross-sectoral group with interdisciplinary representation, engaged end-users, and implemented online delivery via QR code to address some of these potential implementation barriers. Furthermore, we developed an accompanying guide (i.e., ‘help text’) intended to support the use of the leaflet. Our intent was to create a supportive clinical environment with multiple (digital) delivery options, that encourage the use of the *Choosing Wisely* leaflet [19,23,30,39].

Current information resources often lack sufficient details on material development and structure; this heterogeneity, combined with the varying extent of the materials, may influence their adherence and effectiveness [19]. In our study, participants preferred a concise and easy-to-read leaflet. Clinicians emphasized that it should be limited to two pages to enhance usability, clarity, and practicality. People with low back pain underscored the need for the leaflet to present information clearly and comprehensively while being concise and not too long. Key points should be highlighted and not overlooked, ensuring that the leaflet does not overwhelm or discourage engagement with the content. Future research should focus on evaluating the effectiveness of this *Choosing Wisely* leaflet in real-world settings, using validated outcome measures and qualitative approaches. This could involve assessing its usage in clinical settings and its impact on the accuracy of imaging diagnostics, perceived diagnostic uncertainty, improved patient knowledge, beliefs, satisfaction, and adherence to the recommended treatment guidelines.

#### Implications

Healthcare campaigns, such as *Choosing Wisely*, play a vital role in promoting evidence-based decision-making, by promoting evidence-based guidelines, fostering patient engagement, and raising awareness among healthcare professionals at an organizational level [7,34,56]. The *Choosing Wisely* leaflet is portable and could be used by clinicians to remind and reinforce information discussed during clinical encounters to support patients understanding their diagnosis and optimizing the decision-making. It includes pictures recommended to help patients and consumers understand what is being communicated [54]. The *Choosing Wisely* leaflet is not a decision aid, but an informational resource designed to facilitate clinical discussions and inform patients about evidence-based management strategies for low back pain.

The development process facilitated iterative learning and adaptation. Our findings showed the necessity for clarity, simplicity, and relatability in patient educational materials. Through the think-aloud interviews, we implemented significant revisions based on the feedback received, highlighting the importance of involving individuals with lived experience of low back pain in developing patient materials. This fostered a deeper understanding of the rationale behind the *Choosing Wisely* leaflet’s design choices and its anticipated impacts, such as reducing diagnostic uncertainty and enhancing imaging accuracy. By incorporating diverse perspectives and fostering iterative refinement, our methodology may help future research and clinical settings to optimize their delivery of relevant health information to end-users [14]. These efforts must be sustained and adapted over time to ensure that both patients and clinicians are equipped to make informed decisions.

#### Strengths and limitations

We recruited a convenience sample of people with low back pain. These people may not be representative of the target population. Similarly, the median age of the cohort was less than the estimated population prevalence of people with low back pain indicating a potential selection bias [18]. However, by including the perspective of people across the different age groups enhanced the leaflet’s generalisability. One database was deemed appropriate for reviewing and identifying suitable content for the leaflet. The purpose was to create a scientific basis from the literature; therefore, we did not conduct a full systematic review but instead sought evidence-based information from the scientific literature to guide the content of the programme theory and *Choosing Wisely* leaflet. The inclusion of end-users, such as multidisciplinary practice consultants and people with the lived experience of low back pain ensured a comprehensive understanding of the context-specific practical needs and perspectives relevant to the leaflet’s development. This iterative approach allowed for a structured and efficient exchange of ideas, enabling end-users to review the content at their convenience and provide thoughtful, written input with multiple revisions and adaptations. However, further testing in real-world settings is necessary to ensure the leaflets efficacy and to identify potential mechanisms that can support both clinicians and patients in evidence-based decisions.

## Conclusion

We co-created a patient-facing leaflet that is intended to reduce unnecessary imaging in the management of low back pain, across healthcare sectors. The leaflet was developed, and refined, through multiple steps and methods to ensure it aligns with current evidence, local context, and patient needs. This leaflet may support patient care and outcomes by raising awareness about the harms of the use of imaging, such as overdiagnosis and overuse. Real-world implementation and outcome assessment are crucial for understanding the effectiveness, adherence, and potential impact of the *Choosing Wisely* leaflet.

## Institutional Review Board

North Denmark Region Committee on Health Research Ethics determined that this study was exempt from ethical approval because of its non-interventional design. Signed informed consent was obtained from all participants during the think-aloud interviews. All data were stored on a secure server hosted at Aalborg University and North Denmark Region.

## Data availability

Data will be available upon reasonable request to corresponding author. For copies of the *Choosing Wisely* leaflet, see the appendix or contact the corresponding author.

