## Supplementary Material for "Co-creating a *Choosing Wisely* Leaflet Supporting the Reduction of Imaging Usage in Low Back Pain Management - A Multi-Method Study"

**Affiliations**
1. Center for General Practice at Aalborg University, Denmark.

2. Westmead Applied Research Centre, Faculty of Medicine and Health, The University of Sydney, Australia

3. North Denmark Region, Patient pathways and Economy, Aalborg, Denmark.

4. Department of Health Science and Technology, Faculty of Medicine, Aalborg University, Denmark.

5. Department of Physical Therapy and Occupational Therapy, Aalborg University Hospital, Aalborg, Denmark.

### Appendix 1: Literature search:

(("needs"[Title/Abstract] OR "information requirements"[Title/Abstract] OR "personalised information" [Title/Abstract] OR "information preferences"[Title/Abstract] OR "patient information"[All Fields] OR "patient educat*"[All Fields] OR "health literacy"[All Fields] OR "self-care"[All Fields] OR "self-management"[All Fields])) AND (("Back Pain"[MeSH Terms] OR "Back Pain"[tiab] OR "Backache"[tiab] OR "Lumbago"[tiab] OR "Disc Herniation"[tiab] OR "Spinal Fusion"[tiab] OR "Facet Joint"[tiab] OR "Intervertebral Disc"[MeSH Terms] OR "Lumbalgia"[tiab] OR "Sciatica"[tiab] OR "backache"[tiab] OR "spinal pain"[tiab] OR "Spondylosis"[tiab] OR "Slipped Disc"[tiab] OR "Degenerative Disc"[tiab] OR "Spinal Stenosis"[tiab] OR "Displaced Disc"[tiab] OR "Prolapsed Disc"[tiab] OR "Low Back Pain"[MeSH Terms]))

### Appendix 2: Synthesis of inputs from literature search:

##### Literature search

The literature search initially identified 3,018 papers. In total, 34 papers were deemed relevant and included in the development of the programme theory.

**Articles (PubMed ID):**

37020314,36981835,36963709,36804721,36721165,36657216,36641727,36482802,36455471,36332333,36243600,36223377,36198841,35599466,35358311,35150036,34784310,34531223,34373916,34001396,33310794,32776654,31341823,31227280,29574167,29280231,25959985,24001260,23948852,23585633,23073211,21971757,21719235,16688023.

##### Summary of the literature search.

Patients experiencing low back pain needed education and information regarding their diagnosis and an explanation of their symptoms, presented in calming terms tailored to their condition [6,13,17,30]. They expressed a strong desire for clear and consistent information about treatment options and strategies for supported self-management of their problem and pain [6,8,17,26]. Patients needed knowledge about prognosis expectations, including their work ability [4,17,23]. It was important for patients to be involved in their treatment plan and to have their experiences acknowledged and validated, providing reassurance [5,8,9,15,17,30]. Assistance was necessary in understanding the reasons behind whether further investigation, including MRI scans, was warranted or not [5,11,20]. In terms of content and wording, the significance of language was highlighted in some studies where phrases such as "disc bulge", "degeneration", and "arthritis" were associated with poor interpretation of low back pain [5,11,24]. Studies also emphasized the need for personalized, patient-centered approaches in management; information based on the biopsychosocial perspective was suggested to be part of changing the patient's thoughts about low back pain [3,6,8,15,17,23]. These studies underscored the importance of clear, consistent, and personalized information, and effective communication in enhancing the management and outcomes of low back pain [1–6,8–35].

### Appendix 3: Synthesis of inputs from practice consultants:

Purpose: The practice consultants believe that the purpose of the leaflet is to create reassurance for patients and enable them to manage their pain independently. Additionally, the purpose is to reduce the proportion of MRI scans.

Content: The leaflet should contain understandable information about what low back pain is, that it is rarely "dangerous," and that it is rarely structure-specific, which should help reduce misinformation about low back pain.

- It should encourage self-management strategies by providing patients with an understandable explanation of how they can manage their pain in their daily lives, such as rest vs. activity and relevant exercises.
- The leaflet should provide an overview of relevant treatment options available, including options the private sector (e.g., chiropractor and physiotherapist clinics), the municipality, and in hospital settings.
- The leaflet should also include information about which signs patients should react to (i.e., red flags). Regarding whether one should undergo an MRI scan or not, the main messages from the status article in the Danish Medical Journal (Danish: Ugeskrift fra Læger) should be drawn upon [7]

Target population: The leaflet should primarily target citizens with prolonged courses of low back pain who are at risk of being lost in the system (i.e., multiple referrals).

Delivery: The leaflet should be delivered by a healthcare professional who can supplement with verbal information when it is handed out. It can also be available online. It is also suggested that the leaflet could include a QR code linking to videos with additional information.

Scope: The leaflet should be short and easily understandable, with a maximum of two pages to ensure it fits within the consultation with limited time and help maintain the focus on key points.

### Appendix 4: Program theory for Choosing Wisely leaflet - lower back pain in the North Denmark Region

| **Context** | **Activities** | **Mechanisms** | **Short-term outcomes** | **Mid-term outcomes** | **Long-term outcomes** |
| --- | --- | --- | --- | --- | --- |
| Here we consider both primary and secondary target audiences, as well as their characteristics. | Which activities will we initiate to lay the groundwork/foundation for the desired change? | What is the effective mechanism that facilitates/prevents behavior change over time? | What are the short-term goals associated with the use of the brochure? (Patients, Clinicians & others) | What are the medium-term goals for the use of the brochure? | What are the long-term goals/the overarching vision for the project? |
| **Target group (Patients):**  Patients with lower back pain are recipients of the (product) patient brochure and the information therein.  Characteristics:   - Suffering from pain - Need for assessment/clarity - High level of diagnostic uncertainty - Poor understanding/knowledge of lower back pain or treatment options - May be resource-constrained individuals. - May be stressed individuals. - Might believe that scanning can "provide answers" regarding which treatment is the "only/right" one so they can return to a life-as-before. - May perceive scanning (and referral to it) as validation of their pain ("the doctor wouldn't send me for scanning if he/she didn't believe something was wrong"). - Likely unaware or uneasy with the knowledge that back pain can be disabling without being pathological/dangerous, and that there probably won't be relevant findings on a scan.   **Target group (Clinicians)**  The healthcare professional (general practitioner), tasked with diagnosing, treating, selecting, and referring patients (Agent for change).  Characteristics:   - Agent for change. - The one who has an overview and the actual choice regarding referrals for scanning and possible secondary treatment. - Responsible for targeting information to the patient's needs. - Subject to requirements to deliver treatments of high quality. - Likely unaware of the risks of early/unnecessary scanning and/or trained to view MRI scanning as an economic burden for society (something to save on) but a neutral burden for the patient 🡪 may (mistakenly) perceive scanning as a good supplement for an uneasy patient or unclear diagnosis. | Incorporation of a patient brochure with information about MRI scans tailored to the needs of patients and clinicians.  **For Patients**:   - Information about lower back pain and its progression. - Decision support. - Overview of treatment options. - Action-inspiring; what to do/try when the pain is severe. - Perhaps following the same structure as in Credible Explanations (three themes).   **For Clinicians:**   - Brochure and materials with additional information, decision support, and treatment alternatives targeted at clinicians. - Information on procedures regarding referrals. Better alternatives for difficult patients. - Recognition that a brochure is just one tool and that some patients may not be able to read it (health literacy) and/or may not feel comfortable with general information 🡪 Perhaps a "clinician section" that can provide the doctor with background knowledge for the recommendations, including risks of unnecessary recommendations and knowledge about correct referral criteria (as well as possibly rejecting the referral without relevant suspicion, if implemented). | **Mechanisms that facilitate:**  **For Patients:**   - Reassurance = The brochure provides calmness and creates a sense of security. - Patients feel seen and heard. Patient involvement = Patients can read up on lower back pain and become more knowledgeable about their treatment options. - Better understanding of their own situation, illness, and alternatives in the present. - Less worry about having something serious (red flags). - Validation of their experience = The brochure acknowledges the pain condition/experience. Normalization of pain; it's normal for many (20%) to experience pain...   **For Clinicians:**   - Assistance in providing patients with an explanation for why their pain persists and treatment options. - Can save time as the clinician doesn't have to go all the way around the explanation and reasoning. - Assistance in remembering the criteria for/basis for why an MRI scan is given or not given. - Opportunities to encourage patients to wait and see.   **Mechanisms that counteract:**  **For Patients:**   - Patients may be adamant that an MRI scan is the best/only option. - Patients adopt a waiting position until they have their scan. - Patients are not open to the idea that their status and treatment needs may change. - Build trust in the healthcare system and the doctor's treatments (same message distributed across multiple channels).   **For Clinicians:**   - Lack of knowledge about alternatives to scans and what to do with complex patients. - Resistance to change. - Scans become a parking lot. - Scans free up space and create flow. - Some specialists (e.g., neurosurgeons) require imaging diagnostics. - Scans can become outdated. - Lack of trust in other healthcare professionals. - Physician's fear of making mistakes, including defensive medicine. - Practice habits (locked behavior). - Patient demand => We should be able to help the doctor with formulations/arguments against scan-on-demand, including that it is not possible (outside the referral framework).   **Other:**   - How many patients are actually referred elsewhere? | **For Patients:**   - Patients who feel more secure in the present. - Patients who feel heard, seen, and have gained some knowledge from the consultation (+ validation). - Providing patients with an overview of their treatment options in the situation (opening up for new possibilities). - Patients understand the chances of improvement. - Postponing the scan to the future/trying other options first. - Reduced need for MRI scanning. - Reduced diagnostic uncertainty.   **For Clinicians:**   - Avoid scaring patients. - Provides the clinician with a script when informing the patient. - Clinician becomes more knowledgeable about other treatment options available. - Reduces the physician's fear of making mistakes. - Fewer patients returning due to incorrect referrals. - Reduced diagnostic uncertainty.     . | **For Patients:**   - Faster diagnosis 🡪 Avoid unnecessary sick leave and/or loss of function. - Reduce the number of consultations. - Avoid patients feeling like they're being tossed around - they're in a process. - Reduced diagnostic uncertainty.   **For Clinicians:**   - Clinicians become more confident in recommending alternatives to MRI scanning. - Reduced diagnostic uncertainty. | - Reduction in the number of MRI scans for lower back pain. - Reduction in expenses related to MRI scans. - Increased patient satisfaction and trust in the healthcare system. - Fewer patients believing that their back pain is undiagnosed and dangerous, ideally leading to more patients returning to/continuing to work, etc., despite the pain.   **Knowledge for us**:   - Better understanding of how informational material (patient brochures) can be integrated into patient treatment. - Identify/create better understanding of the mechanisms through which the brochure can support behavior changes. - Identify information needs among doctors, patients, etc. |

### Appendix 5: Interview-guide

In a moment, you will be handed a leaflet containing information about back pain. I would like you to read the leaflet. It is important to emphasize that this is not a test – I am simply interested in hearing your thoughts on the content and the text. During this exercise, I would like you to 'think out loud' while reading and looking at the content. This means that you should not only read the leaflet but also share your thoughts while doing so, and preferably express the impressions you get when looking at the content. I would like you to share your thoughts, uncertainties, confusion, and questions. You can also ask me questions along the way. The questions can be about what you see, think, or do. The exercise will take a maximum of 20 minutes.

You are welcome to read the text aloud if it helps, but it is not necessary if you prefer to omit it. It is best if you imagine that I am not present at all. I will take notes along the way while you 'think out loud'; I do this to ensure that we remember everything. However, if you are silent for a long time, I may interrupt your reading and ask about what you are thinking. Remember, there are no 'wrong' or 'bad' comments. It is very important that you are completely honest when thinking out loud – there is no need to try to be polite. We are interested in your honest opinions. You are here to help us understand whether our leaflet is useful or not. If there are things that confuse you or you do not understand, it is very likely that others will face the same challenges. Please note that I am recording today's session, but everything you say and do will be treated confidentially, and you will remain anonymous.

At first, it may seem a bit strange to think out loud since it is not something people do often. However, this is completely normal, and most people quickly get used to it. Do you have any questions before we start?

| **Focus** | **Questions** |
| --- | --- |
| **Examples of probing questions (during interview).** | Probing questions in a think-aloud exercise are designed to encourage participants to verbalize their thoughts in a neutral manner while working on a task or problem – in this case, a leaflet. Ask participants to elaborate on their thought process or provide more details about what they are currently thinking or doing.   - "Can you explain what you're thinking right now?" - "How does this information affect your thoughts?" - "You're not saying anything right now?" - "Can you explain your thought process?" - "I can see you're pausing." - "Where in the text are you right now?" - "What are your thoughts on this section?" |
| **Communication and Formats *(after the participant has read through the*** leaflet***)*** | - "How do you think this information could be best conveyed – should it be something other than the paper leaflet you've seen today?" - If no answer, mention examples such as: video, digital format, posters, audio, etc. Ask why or why not. - "How would you adapt this leaflet for different platforms, such as online marketing or digital distribution?" - "How would you tailor the content and design to appeal to individuals with back pain?" - "What was your initial reaction to the leaflet?" - "What criteria do you use to assess whether the information in the leaflet is useful?" - "How would the information be strong and compelling for individuals experiencing back pain?" |
| **Debriefing** | - "Now that you've read the entire leaflet, what do you think its purpose is?" - "What does the leaflet aim to convey to the reader?" - "Can you describe how you understand back pain?" - "What considerations do you have in mind to ensure that the leaflet is easy to read and understand?" - "Where did you find the leaflet difficult to understand?" - "Can you tell me about your overall impression of the leaflet?" - "What were the primary challenges?" - "What additional elements or improvements would you consider adding to the leaflet?" - "The overarching goal is to reduce the need for scans - How would you evaluate this leaflet in terms of achieving that goal?" - **Finish with notes from your pad.** |

### Appendix 6: REDCap questionnaire

| Thank you for your participation! |
| --- |

Over the past week - how have your worst lower back pains been on a scale from 0 - 100?

0 = no pain	100 = worst imaginable pain

*(Place a mark on the scale above)*

Over the past week - how have your average lower back pains been on a scale from 0 - 100?

0 = no pain	100 = worst imaginable pain

*(Place a mark on the scale above)*

How long have you lived with your low back pain (years/months)?	__________________________________

Do you experience pain anywhere other than your lower back?

Har du ondt andre steder end lænderyggen?

Yes

No

Do you have pain anywhere other than your lower back?

- Back (including neck pain)
- Stomach
- Shoulder
- Chest
- Upper arm
- Lower arm
- Elbow
- Wrist
- Hand
- Hip
- Pelvis
- Groin
- Thigh
- Knee
- Shin
- Calf
- Ankle
- Foot
- Headache
- Widespread pain (e.g., fibromyalgia/widespread pain)

Gender

Male

Female

Other

In what year were you born?

__________________________________

Do you engage in sports/physical activity?

Yes

No

How many hours per week do you engage in sports or physical activity on average?

__________________________________

Do you take pain medication?

Yes

No

Prefer not to answer

How often do you take pain medication?

Rarely

Once a month

Once a week

More than once a week

Daily

Prefer not to answer
